# Feasibility, Acceptability, and Cost of Community-Based Self-monitoring among Sex Workers Testing Positive for COVID-19 in Zimbabwe: A Mixed-methods Study

**DOI:** 10.64898/2026.03.16.26348020

**Authors:** Itai Kabonga, Collin Mangenah, Constancia Watadzaushe, Claudius Madanhire, Nancy Ruhode, Yasmin Dunkley, Karin Hatzold, Elizabeth Corbett, Frances M Cowan, Euphemia L Sibanda

**Author notes:** Corresponding author, Itai Kabonga, Centre for Sexual Health and HIV/AIDS Research- Zimbabwe, 10 Coronation Road, Greendale, Harare, Zimbabwe.

## Abstract

**Background:** Sex workers struggled to adhere to isolation guidelines following COVID-19 diagnosis because of financial pressure to keep working. We co-developed and evaluated for feasibility, acceptability, and cost an intervention for promoting isolation and community-based self-monitoring for COVID-19.

**Methods:** Sex workers testing positive for COVID-19 received the following co-developed intervention: i) risk-differentiated support, including immediate hospitalization and/or treatment for serious illness, and community-based self-monitoring for those at risk of progressing to severe illness, ii) food packs lasting two weeks. Using Proctor’s Framework, we interviewed purposively selected health-workers and sex workers before intervention implementation (26 sex workers and 24 health workers) and during implementation (8 sex workers of whom 5 tested positive, and 5 health workers) to evaluate the intervention. We determined intervention development and implementation costs using program data.

**Results:** The intervention was implemented between March-June 2023. Sex workers and health workers reported that the intervention was highly acceptable and was implemented with fidelity. Food packs were highly appreciated; participants said they promoted isolation although vulnerability to non-food financial pressures persisted. Unanticipated impacts were increased testing uptake following introduction of food packs. Self-monitoring at home was acceptable although fear of stigma prevented some participants from seeking the needed support. The cost per sex worker testing positive was $49 and $54 respectively excluding/including intervention co-development costs.

**Conclusion:** A co-developed intervention for promoting isolation and community-based self-monitoring for COVID-19 was feasible and acceptable, with costs comparing favorably with similar interventions. Addressing stigma could optimise implementation and potential for future pandemics.

## 1. Introduction

COVID-19 caused substantial mortality and morbidity when it emerged in December 2019, with over 770 million infections and 6 million deaths reported cumulatively worldwide by September 2023 [1]. Testing, isolation of those testing positive and contact tracing were key to reducing the spread of the disease [2, 3]. At the height of COVID-19 outbreaks in Zimbabwe and similar settings, uptake of COVID-19 testing was sub-optimal, with challenges at the level of the health system, community, and patient. Health system barriers included supply-chain challenges for test kits, shortage of trained personnel, particularly when only nucleic acid amplification tests were the standard of care, with long turn-around times which limited the utility of the results [4]. Community-level factors included stigma associated with COVID-19 testing and myths surrounding the disease [5]. Patient level factors included challenges accessing testing centers, which was worsened by lockdowns and fear of negative impacts of testing such as requirement to isolate, or stigma [3]. These factors increased the vulnerability of communities to COVID-19.

Sex workers were particularly vulnerable to COVID-19 because the nature of their work exposed them to COVID-19 acquisition from their clients, and yet lockdowns and requirement to isolate (when diagnosed positive) meant that they could no longer earn a living [6, 7]. In Zimbabwe and similar settings, sex workers have no access to social protection interventions and COVID-19 further marginalized them [8]. Development of a COVID-19 testing model that was suitable for sex workers was critical for reducing their vulnerability. The Key Populations (KP Program) in Zimbabwe provides HIV and sexual and reproductive health services (SRH) for sex workers across the country [9]. To ensure access to COVID-19 testing among sex workers and prevent clinic level transmission (important for preventing closure of the clinics) COVID-19 testing was integrated within these HIV and SRH services. Individuals visiting the clinics were required to undergo symptom screening (and testing if symptom screened positive) before they could access the clinic.

To address the unique barriers that sex workers faced in relation to COVID-19, we co-developed an intervention for providing COVID-19 testing and post-test support services among sex workers who were enrolled in the KP Program. We aimed to integrate post-test support with standard protocols of risk-differentiated community-led services employed within the KP Program. Risk differentiated support and management of people with COVID-19 is supported by the literature [10] - mild cases can be managed outside health facilities through implementation of strategies such as telemedicine (use of telecommunication technologies to diagnose, treat and offer health care support), with severe cases requiring hospital management. We describe the intervention development, and investigate implementation success, including fidelity, feasibility, and acceptability, as well as the cost of intervention development and implementation.

## 2. Methods

### 2.1. Study Design

Working with sex workers, health workers providing health services to sex workers, sex worker peer educators and relevant stakeholders, we co-developed an intervention for facilitating community-based self-monitoring for COVID-19 among sex workers. We piloted the co-developed intervention between March to June 2023 at 12 clinics in the KP program and evaluated it using mixed methods including analysis of program data, qualitative studies and costing studies.

### 2.2 Setting: The Key Populations Program in Zimbabwe

The Key Populations program is a national program that provides sexual and reproductive health, and HIV prevention and care services for sex workers (female, male and transgender). Established in 2009, the program has national coverage with 86 implementation sites in 2023, comprising 12 static local sites, 34 local mobile clinic sites, 10 Drop-in Centres and 30 highway mobile clinic sites. Over 30,000 sex workers were seen in the program in 2020, representing 75% of the estimated population size of 40,000. Uptake of services within the program is documented using electronic tools. Implementation of the KP program is supported by a network of peer educators called microplanners (sex workers enrolled in the KP Program who are trained to provide health education and risk-differentiated support to fellow sex workers for continued engagement in the program and adherence to HIV, STI and other disease prevention/treatment interventions). Microplanners are supported by outreach workers – program employees who are responsible for supervising recruitment and retention of sex workers.

From the beginning of the COVID-19 pandemic to June 2023, the program operated COVID-19 symptom screening procedures among attendees before they entered the clinic waiting area. This included temperature checks and administering a short questionnaire on COVID-19 symptoms. Sex workers who symptom screened positive (i.e., were likely to have COVID-19) were offered testing, see symptom screening questionnaire in supplement (**S1**).

### 2.3 Intervention Development and Summary of Agreed Intervention

We held a workshop with sex worker representatives including female sex workers, men having sex with men and transgender women in October 2021 to codevelop a model for providing COVID-19 testing and community-based follow up of sex workers testing positive for COVID-19. In attendance were 26 people including eight microplanners/peer educators (seven female, one male), eight sex workers (six female and two transgender), four clinic-based program staff, three program managers and three study staff working on the project. The meeting (a group discussion) was facilitated by one of the program managers as is the norm when meetings are held with sex worker representatives. Discussions followed a guide that explored challenges to adherence to isolation guidelines and potential solutions. In this workshop, sex workers reported that it was difficult to comply with the requirement to isolate following COVID-19 diagnosis because they could not afford to stop working. Workshop attendees agreed that a food hamper could help relieve financial pressures and enhance adherence to isolation guidelines. We collaboratively agreed on a food hamper containing basic food items adequate for a family of four over a 14-day period, valued at US$25. In addition, workshop attendees agreed on risk-differentiated support for sex workers diagnosed with COVID-19, (**Figure 1**).

**Figure 1:**
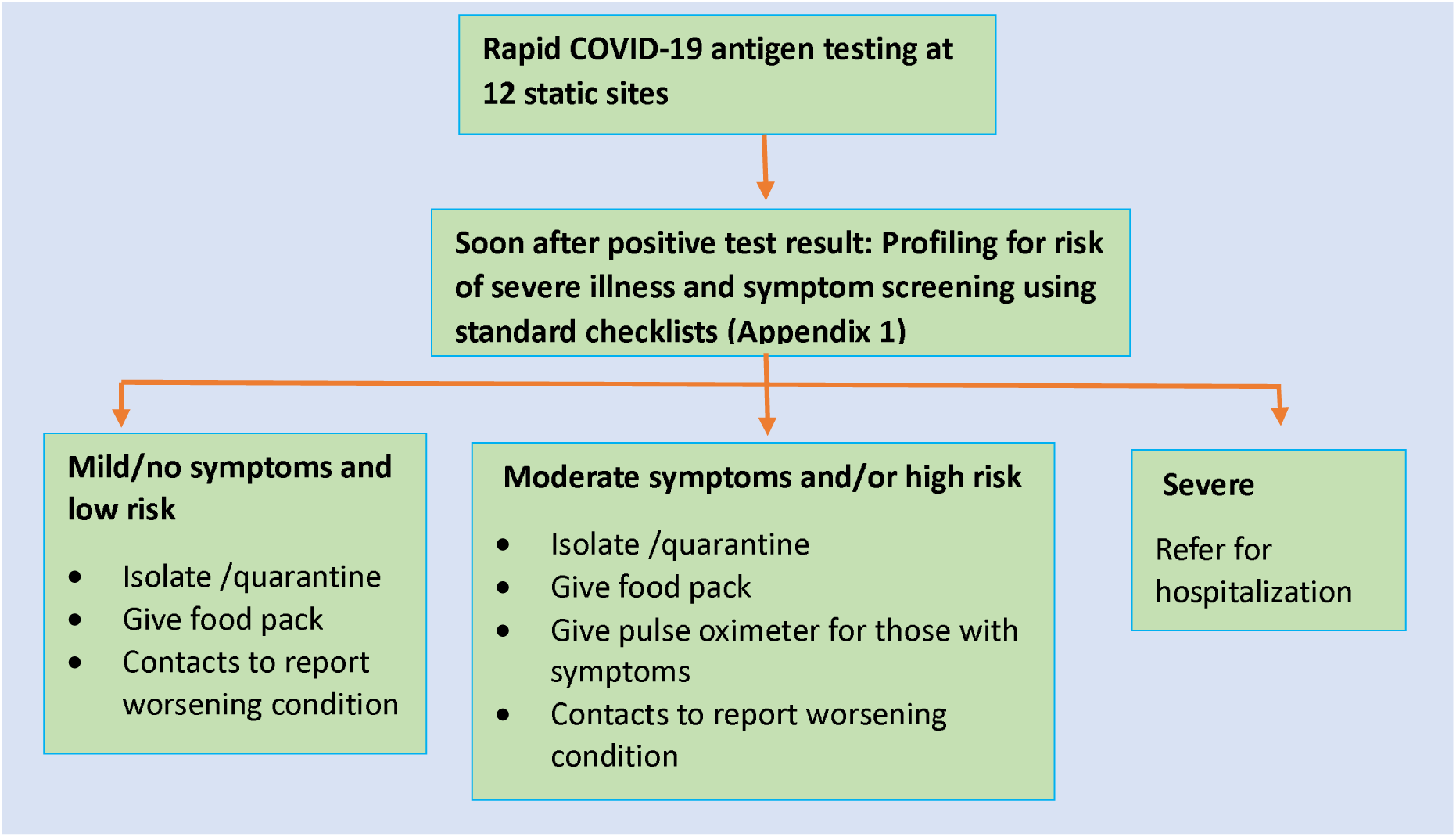
Summary of Post-test Support for Sex Workers Testing Positive for COVID-19.

Upon COVID-19 diagnosis, a sex worker was screened by a health worker for severity of current symptoms and potential to suffer from serious illness (**S2 and S3 for the screening checklists**). Participants with serious illness would be referred for immediate hospitalization and/or treatment, while those who had moderate symptoms and/or risk of developing severe illness were assigned to community-based self-monitoring and support of food pack to promote isolation and contacts of health workers to call in case of worsening illness (Figure 1). Participants doing self-monitoring also received pulse oximeters and instructions on how to use them, interpret results and what actions to take if readings indicated worsening disease. Participants were told that if their oxygen saturation level is above 95% indicating mild illness, they needed not to be alarmed but continue isolation and self-monitoring. Participants were also advised to continue isolating and self-monitoring if oxygen saturation levels were greater than 90% but less than or equal to 95% as this indicated moderate illness. Participants were advised to seek support for travel to hospital if symptoms worsened and oxygen saturation levels were less than 90% by either contacting the program directly or their microplanners, Table 1. The agreed intervention was implemented between March to June 2023 at 12 static clinics across Zimbabwe.

**Table 1:** Categorization of illness and steps to take during self-isolation.

| Category of illness | Indicators | Action to take |
| --- | --- | --- |
| Mild | Oxygen saturation level above 95%<br>No signs of respiratory distress | Home isolation<br>Continue self-monitoring |
| Moderate | Oxygen saturation is greater than 90% but less than or equal to 95%<br>Weak but can walk | Home isolation<br>Continue self-monitoring |
| Severe | Oxygen saturation is lower than 90%<br>Patient is too weak to walk | Notify health workers for support with immediate hospitalization. |

### 2.4 Evaluation of the Intervention

Evaluation of the intervention was done according to Proctor’s Framework [11], with a focus on the following outcomes: acceptability, feasibility, appropriateness, adoption (uptake), fidelity, and costs. Below we detail the methods that we used to determine each outcome.

#### 2.4.1 Determining Adoption

##### Program Data Collection

Program data documenting adoption (uptake) of the intervention were collected by program staff using tablets, documenting offer and uptake of screening for COVID-19 symptoms, screening outcomes, uptake of COVID-19 tests (for those screening positive) and corresponding results. Limited demographic information was collected for clients accessing the services including gender, date of birth, and age.

##### Sampling and Analysis of Program Data

We used data for all participants who were screened during the study period. Data were analyzed descriptively using STATA version 14. We summed up the numbers of sex workers screened and tested before and during the intervention, and computed weekly numbers screened, tested, and results for the same periods.

We used the program data to identify eligible respondents for purposive sampling for the qualitative study, determining uptake of intervention components and confirmation of intervention fidelity i.e. completion of risk assessments and implementation of risk differentiated support.

#### 2.4.2 Determination of Acceptability, Feasibility, Appropriateness, and Fidelity

##### In-depth Interviews with Sex Workers

In-depth interviews were conducted with sex workers from 12 static clinics in Mbare (Harare), Bulawayo, Gweru, Masvingo, Beitbridge, Mutare, Forbes, Karoi, Chinhoyi, Victoria Falls, Kadoma and Chirundu to explore the acceptability, feasibility, appropriateness, and fidelity of the intervention. Eligible sex workers accessed HIV and sexual reproductive health services from the KP program, with purposive sampling to include sex workers testing either negative, positive, and representation by age and clinic type (those in large/small cities). Interviews were conducted in two phases: 26 interviews before self-monitoring intervention implementation, capturing hypothetical acceptability and feasibility, and 8 interviews during intervention implementation, capturing actual acceptability, appropriateness, feasibility, and fidelity. Eligibility for participation in phase two interviews was based on prior community-based self-monitoring, regardless of involvement in phase one interviews. Interviews were conducted by experienced social scientists in languages preferred by participants using guides that elicited views on the intervention, experiences of the intervention, what worked well, what worked less well and suggestions for improvement. Interviews lasted between 40 minutes to one hour and were audio recorded.

##### In-depth Interviews with Health Workers

In depth interviews were conducted with 24 health workers before the self-monitoring intervention and 5 health workers during the self-monitoring intervention. Only health workers involved in supervising community-based self-monitoring were interviewed regardless of participation in phase one interviews. Purposive sampling was done to include nurses, outreach workers and peer educators who implemented the intervention for at least two months. Interviews were conducted by experienced social scientists in languages preferred by participants using guides that explored views on the intervention, experiences of implementing the intervention, what worked well, what worked less well and suggestions for improvement. Interviews lasted between 40 minutes to one hour and were audio recorded.

##### Data Handling and Analysis

Thematic analysis was used to analyze qualitative data [12]. In-depth interviews were transcribed verbatim by trained research assistants. Qualitative data analysis started as soon as data collection began. Within two days of each interview, detailed field notes were written with attention to emerging themes. We further immersed ourselves in the data by reading transcripts to identify patterns in the data. Weekly data analysis meetings were conducted among social science researchers who were involved in data collection to explore emerging themes. The discussions focused on themes related to acceptability, feasibility, appropriateness, and fidelity. During these meetings, emerging themes were either merged or unbundled depending on supporting data. Following transcription, analytic summaries that drew comparisons within and across participants were written for each interview, followed by construction of a coding framework inductively with major codes covering accessibility to COVID-19 services, attitudes and perceptions to COVID-19 services and behavioural response to COVID-19 services (**S**4). Coding was done using Nvivo version 14[13] by one coder with consensus meetings held weekly to review coding until the team determined that further discussions were not necessary.

#### 2.4.3. Determining Costs

Full annual economic costs of implementing the community based self-monitoring intervention were estimated using an ingredients-based micro-costing approach [14, 15].Costs of the intervention included start-up co-development and implementation.

##### Cost Data Collection

Detailed resource utilization data were collated between March and June 2023 alongside implementation of the community based self-monitoring intervention taking a health system perspective [16, 17]. This ensured that we were able to prospectively observe and get a full understanding of how intervention activities were implemented and to collect costs in real-time. Financial expenditure records, invoices, and receipts provided by the implementing organization were used as a source of cost estimates.

###### a. Co-development Costs

Co-development costs included teas, lunches, and transport reimbursements for workshop participants. Start-up co-development activities were considered capital costs with a useful life lasting more than one year [18]. Amortisation was therefore applied over a four-year economic life and at a 3% discount rate.

###### b. Costs of Intervention Implementation

Implementation costs comprised personnel time, the initial COVID-19 test, the pulse oximeter equipment, medical prescriptions and masks, and non-medical supplies such as food packs, communications and transport.

Personnel costs were estimated based on health provider reported time required for consultation, treatment, referral, and follow-up (including any telephone calls) of both mild and any severe cases [19, 20]. Personnel salary costs were used to estimate an hourly rate and then combined with the average duration of each activity (treatment, referral, or follow-up).

Although costing only $8 per unit, in this study the pulse oximeters for moderate cases were classified as capital equipment costs rather than a supply input (anything less than US$100 or not lasting more than a year). This was based on the life span of a pulse oximeter which ranges between five and seven years, although the SpO_2_ sensors lasts no more than 1 to 1.5 years with regular usage [21]. A two-year economic life was therefore considered appropriate and discounting at a 3% discount rate applied [18].

Medical supplies included the initial COVID-19 test, prescription medication, and K95/N95 masks (medical) for moderate and severe cases. Non-medical supplies comprised of food packs for a family of four allotted to both moderate and severe cases. Programme costs also include other costs for communications and transport related to following up severe cases by medical staff, and transport to ferry severe cases to hospital.

##### Cost Data Analysis

Costs of start-up co-development and intervention implementation were summed up to estimate total programme costs separately for development and implementation and for implementation alone. As we took a health system perspective our analysis focussed only on the costs incurred by the provider (Ministry of Health in a scale-up scenario) although a broader societal perspective, including out of pocket costs (transport and food) and productivity losses borne by patients would have helped capture the full costs including those that discourage patient uptake of health services. Ultimately, we divided these total programme costs by the number of positive cases to estimate cost per participant self-monitoring. Costs are tabulated by input type and by level of severity of illness in 2023 United States dollars.

### 2.5 Ethical Considerations

The study was approved by Medical Research Council of Zimbabwe (MRCZ), ref MRCZ A2872, London School of Hygiene and Tropical Medicine (LSTM), ref 26931 and the WHO Ethics Review Committee (ERC), ref CERC.0160. Participation in the study was voluntary, with written informed consent for all qualitative study participants.

## 3. Results

Intervention development was done in October 2021, followed by the pilot and costing study between March to June 2023. We present themes on adoption (uptake of testing), acceptability, feasibility, appropriateness and fidelity to risk assessment and risk differentiated support, challenges posed by requirement to isolate after COVID-19 diagnosis and acceptability of the food pack, feasibility, acceptability, and fidelity to self-monitoring at home.

### 3.1 Adoption (Uptake of Testing)

We describe uptake of COVID-19 testing during the period four months before intervention (November 2022-February 2023) implementation and the four months during implementation (March -June 2023). During the four months before the intervention 8,658 sex workers were screened for COVID-19 (monthly range:1,384-2,894), of whom 214 (2.5%) met criteria for testing and 105 (49.1%) were tested. This compared with 9,519 screened during intervention implementation (monthly range: 2,287-2536) of whom 433 (4.5%) met criteria for testing, and 282 (65.1%) were tested. Introduction of the intervention (March to June 2023) was associated with increases in the weekly number of COVID-19 tests: 5 tests were done per week (0-24 range) before the intervention, compared with 15 tests per week (2-37 range) during the four months of the intervention. **Figure 2** shows the weekly number of sex workers screened and tested four months before intervention implementation and during the four months of intervention implementation.

**Figure 2:**
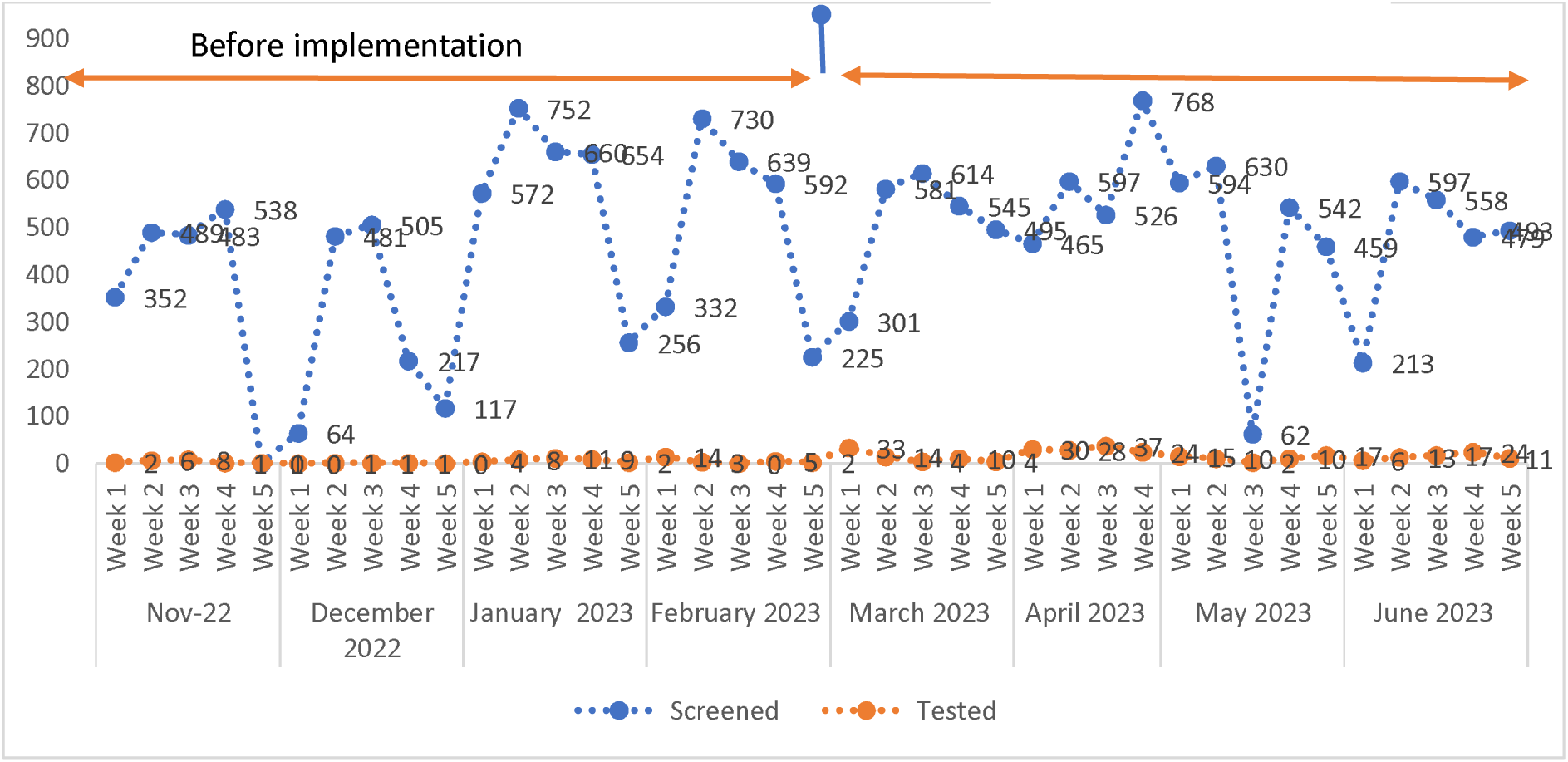
Weekly number of sex workers screened and tested before and during intervention implementation.

### 3.2 Acceptability, Feasibility, Appropriateness, and Fidelity of the Intervention

Prior to community based self-monitoring intervention, we interviewed 26 sex workers and 24 health workers. During the intervention, we interviewed 8 sex workers and 5 health workers. Characteristics of interviewed sex workers and health workers are shown in **Table 2**. Sex worker ages ranged from 20-57 years (mean 33 years); 33 were female with one other. All health workers were female, with ages ranging from 23-62 years and a mean age of 38 years.

**Table 2:** Participant Characteristics of Sex Workers and Health Workers.

| Characteristic | Sex Workers (N/%) | Health Workers (N/%) |
| --- | --- | --- |
| **Age** |  |  |
| 20-29 | 14(41.2) | 5(17.2) |
| 30-39 | 12(35.3) | 10(34.5) |
| 40-49 | 6(17.6) | 6(20.7) |
| 50-59 | 2(5.9) | 5(17.2) |
| 60-69 | - | 2(6.9) |
| Missing | - | 1(3.4) |
| **Gender** |  |  |
| Female | 33(97.1) | 27(93.1) |
| Male | - | 1(3.4) |
| *Other | 1(2.9) | 1(3.4) |
| **Marital Status** |  |  |
| Single | 17(50) | 10(34.5) |
| Married | 5(14.7) | 12(41.4) |
| Separated | 2(5.9) | - |
| Divorced | 8(23.5) | 1(3.4) |
| Widowed | 2(5.9) | 5(17.2) |
| Missing | - | 1(3.4) |
| **Education Levels** |  |  |
| Primary | 5(14.7) | - |
| Secondary | 26(76.5) | 6(20.7) |
| Tertiary | 3(8.8) | 22(75.9) |
| Missing | - | 1(3.4) |
| **Occupation** |  |  |
| Sex work only | 16(47.1) | - |
| Sex work and other | 18(52.9) | - |
| Nurse | - | 16(55.2) |
| Outreach worker | - | 11(37.9) |
| Other | - | 2(6.9) |
| **Type of Test** |  |  |
| Professional | 24(70.6) | - |
| Self-test | 5(14.7) | - |
| Not tested | 5(14.7) | - |
| **Period with Organisation** |  |  |
| Greater than 5 years | - | 14(48.3) |
| Less than 5 years | - | 14(48.3) |
| Missing | - | 1(3.4) |
| **Period at Current Position** |  |  |
| Greater than 5 years | - | 8(27.6) |
| Less than 5 years | - | 20(69) |
| Missing | - | 1(3.4) |
| **Total** | **34(100)** | **29(100)** |

\*Other on gender denotes individuals who identified themselves as gay

### 3.3 Acceptability, Feasibility, Appropriateness and Fidelity to Risk Assessment and Risk Differentiated Support

Evidence from the program electronic data showed that risk assessment and risk differentiated support protocols were followed by health workers for all nine participants who tested positive during the intervention. Health workers reported no challenges in conducting risk assessment, stating that the tools were appropriate and easy to complete. Program data showed that of the 9 positive cases, 8 had low vulnerability to serious illness, figure 3, with one being vulnerable. The participant who was vulnerable received the appropriate commodities and support for community-based self-monitoring.

**Figure 3:**
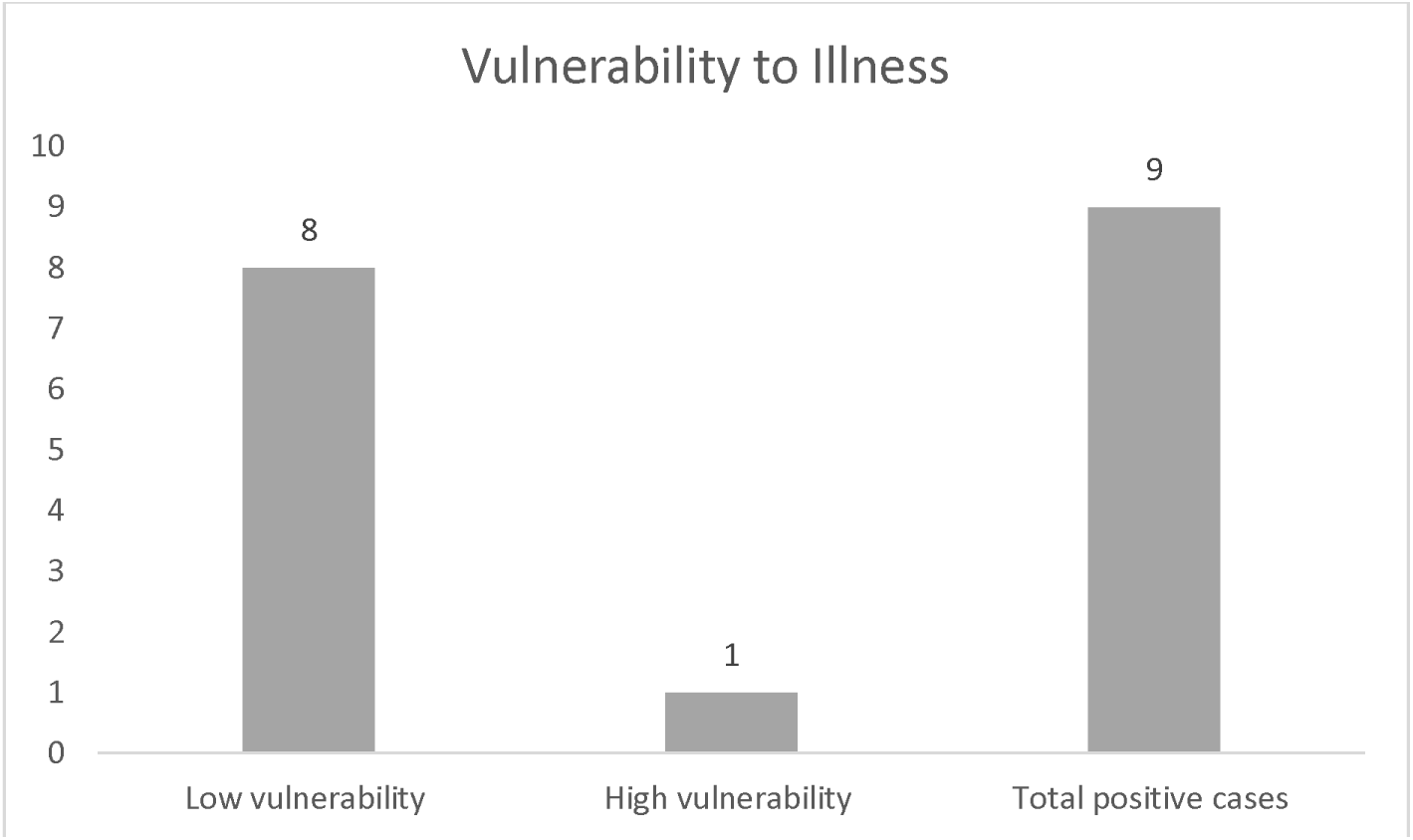
Showing Vulnerability of Cases to Severe Illness.

Interviewed health workers also reported that assessment of disease severity was done without challenges. Health workers captured disease severity electronically, and one participant was found to have severe illness and was assisted to go to a hospital for further care. According to study protocol, participants with severe illness required immediate hospitalization. Five sex workers had mild symptoms, and three had moderate symptoms. They were all managed according to **Figure 1** above, encouraged to isolate, supported with food packs, pulse oximeters and contacts of health workers, showing high fidelity to the intervention. Interviews with clients confirmed that the risk assessments and differentiated support were done:

> “… *I was inserted* (a pulse oximeter*) during testing for COVID* (by a health worker to measure oxygen saturation)…” (Female Sex Worker [FSW], aged 40 to 49 years).

Regarding hospitalisation support, a participant who had been severely ill confirmed getting help from a nurse to go to a nearby hospital for treatment. Participants with mild or moderate symptoms reported being provided with treatment to relieve symptoms. This was appreciated by clients who contrasted with public sector clinics where treatment was sometimes not available “*… like at the clinics* (public sector clinics) *nowadays… you can be told that you have COVID, but they can just send you back home…without giving you anything or offering you any help* (treatment)” (FSW, aged 20 to 29 years).

### 3.4 Challenges Posed by Requirement to Isolate after COVID-19 Diagnosis

Both sex workers and health workers interviewed prior and during the intervention reported that if sex workers did not have food or material support it was difficult for them to isolate after testing positive – they were forced to keep working to earn food for the family. Before implementation of the intervention, there were reports that many sex workers refused the offer to test because they worried about being told to isolate if they tested positive:

> *“If you are told you cannot leave the house (because of COVID-19 diagnosis) …how then do I survive?… so if you tell me to stay indoors what will I be eating?”* (FSW, aged 30 to 39 years).

> *“…because mostly they say, do I eat soil if I isolate?”* (Health Worker [HW], aged 50 to 59 years).

### 3.5 Acceptability of the Food Pack

Prior to the intervention, health workers and sex workers alike reported that the food pack would be acceptable for supporting adherence to isolation. Vulnerability to food insecurity was identified by both as an impediment to adherence to isolation. During the intervention all but one sex workers found the food pack acceptable and effective for facilitating isolation. They all understood the purpose of the food pack was to promote adherence to isolation. Program data shows the disbursement of food packs to all eight eligible participants (those who tested positive for COVID-19 with mild or moderate symptoms). It appeared that at the time they received the food packs it was a pleasant surprise as they had not been expecting it *“No one will be unhappy on things he or she was not expecting… I will not be under pressure because of COVID*” (HW, aged 50 to 59 years). Both sex workers and health workers had consensus that food packs helped improve adherence to isolation:

> *“Yes, it helped me a lot because if I had fallen sick without food in the house… or if I had nothing to give to my child, I would have gone out to look for food… but this helped me not to go out”* (FSW, aged 20 to 29 years).

> *“… she was given a food hamper with 10kgs mealie meal, kapenta, cooking oil… Ehh, that really helped… I also think that made her continue with the isolation.”* (HW, aged 40 to 49 years).

There was evidence of unanticipated effect of the food packs, with health workers reporting an increase in numbers of sex workers symptom-screening positive. Health workers said they suspected some sex workers who would previously give false responses during symptom screening felt more comfortable to report symptoms as they no longer feared isolation given the offer of a food packs for those with COVID-19. The unanticipated increase was reported after word spread in the communities of the offer of food packs to sex workers testing positive. Program data indicate a steady increase in the number of sex workers tested during phase two, coinciding with the distribution of food packs. Health workers reported that some sex workers who reported symptoms were disappointed when they did not test positive as this excluded them from the food pack:

> *“At some point there was pressure that anyone who thought they had flue wanted to just test so I think because of the food hamper some thought no, the result is false because I am sick with flue. Everyone thought if I test, I should get a positive result and leave with a food hamper”* (HW, aged 40 to 49 years).

Although the food packs were helpful for promoting adherence to isolation, both sex workers and health workers reported that there was residual vulnerability because food packs did not address non-food related financial pressures. Examples of these were rental costs, school fees, or medical bills for the family. Residual vulnerability was not reported before implementation and came out strong during the implementation. There was a suggestion for including direct cash transfer in the package to enhance adherence. There were also isolated suggestions to increase the number of food items that were in the food pack:

> *“So, for one to be able to look after themselves in the house for 10 days, 14 days they cannot because they will be getting pressure from the owner of the lodge, I want my money I want my money I want my money”* (FSW, aged 20 to 29 years).

> *“…if only they can add rice or spaghetti to cater for the afternoon”* (FSW, aged 40 to 49 years).

### 3.6 Feasibility, Acceptability, and Fidelity to Self-monitoring at Home

Reports from sex workers showed that they had found it feasible to self-monitor for severity of their illness at home using pulse oximeters and other indicators of worsening illness (difficulty breathing, fast breathing, chest pains, difficulty walking and confused thinking). All but one clearly and correctly described how they used the pulse oximeter to self-monitor:

> *“…(the nurses said) if you observe a low reading you can call, if you are not free to come here, you can visit any local clinic”* (FSW, aged 30 to 39 years).

There was evidence that participants could benefit from reminders of how to use the oximeter (**S5**). For instance, a sex worker aged 20 to 29 years was worried that her oxygen saturation numbers were going up when in fact it was a good thing. This was despite having received both verbal and written instructions (including in local languages) on interpretation of pulse oximeter readings.

There was evidence that participants did not always seek help when they found that their illness worsened. Some participants reported feeling worse, and that they could have benefitted from treatment and care, but they did not report or seek care because they were afraid of potential negative impact. Such participants worried that contact/visits from the program team could cause inadvertent disclosure of the illness to landlords or neighbors, which could cause problems:

> *“…I was afraid that if the landlord gets to know about it, I could be served with an eviction notice, yet I did not have anywhere else to go”* (FSW, aged 20 to 29 years).

Part of self-monitoring at home encompasses preventing the spread of COVID-19 in the household. However, many sex workers live in crowded conditions which makes it very difficult for them to prevent the spread of COVID-19. Sex workers reported taking their children to relatives, neighbors, and friends to avoid exposing them to COVID-19. A respondent submitted “*when I got home, … I said to the landlord’s grandchild…, please sleep with my child*” (FSW, aged 20 to 29 years). This was also confirmed by an interviewed health worker at the site where the client tested positive. Other strategies to help the situation included making sure the children stay away from the client although living in the same room “…*then I move others* (children) *from the bed to sleep on the floor so that I do not infect them with covid*” (HW, aged 40 to 49 years).

### 3.7 Costs of Intervention Co-development and Implementation

Table 3 shows the programme and unit costs of intervention co-development and implementation classified by input type. The annualized cost of co-development for the intervention was $43. The annual total cost of providing the actual intervention to promote adherence to isolation and community-based self-monitoring was $442. Including the cost of development resulted in a total cost of $484. The cost per positive sex worker self-monitoring was $49 (excluding costs of co-development) and $54 when including costs of co-development. Key cost contributors were the non-medical supplies comprising the food packs (41%) for both mild and moderate cases, the initial COVID test (26%) and medical prescriptions (21%) for both moderate and severe cases. Start-up, personnel time and equipment (pulse oximeters) contributed 9%, 5% and 4% to total costs respectively.

**Table 3:** Programme and Unit Costs of Intervention Development and Implementation by Input Type.

| Phase and cost input type | Activity | Input costs (USD) | Total intervention & unit costs (USD) | % cost contribution |
| --- | --- | --- | --- | --- |
| Startup costs | Inception workshop |  |  |  |
| | Tea | \$20.00 | | |
| | Lunches | \$50.00 | | |
| | Transport Reimbursements | \$100.00 | | |
| **Total costs (Startup)** | | | **\$170.00** | |
| **Annualized start-up cost** | | | \$42.50 | 9% |
| **Equipment costs** | Pulse oximeter for moderate cases | \$20.90 | | 4% |
| **Personnel** | Consultation time for mild cases | \$12.46 | | |
| | Follow-up call (after 2 weeks) for mild cases | \$4.00 | | |
| | Consultation time for moderate cases | \$7.48 | | |
| | Referral time to hospital for severe cases | \$1.25 | | |
| **Total costs (Personnel)** | | | **\$25.18** | 5% |
| **COVID test costs** | Initial test | \$125.27 | \$125.27 | 26% |
| **Medical supplies** | Medical prescription for moderate cases | \$15.00 | | |
| | Mask (K95/N95) for moderate cases | \$31.20 | | |
| | Medical Prescription for severe cases | \$10.00 | | |
| | Mask (K95/N95) for severe cases | \$10.40 | | |
| **Total costs (Medical supplies)** | | | **\$66.60** | 14% |
| **Other supplies** | Food pack for mild cases | \$125.00 | | |
| | Food pack for moderate cases | \$75.00 | | |
| **Total costs (Other supplies)** | | | **\$200.00** | 41% |
| **Other (communications & transport) - severe cases** | Follow-up call (after 2 weeks) for 5 mins | \$0.80 | | |
| **Transportation** | Transportation to hospital for severe cases | \$2.81 | | |
| **Total costs (Other & communications)** | | | **\$3.61** | 1% |
| **Total programme costs** | | | **\$441.56** | |
| Total positive cases |  |  | 9 |  |
| Cost per participant self-monitoring | | | \$49.06 | |
| **Total programme costs (including development & implementation costs)** | | | **\$484.06** | **100%** |
| Cost per participant self-monitoring | | | \$53.78 | |

Table 3 shows the same programme and unit costs when classified by level of severity of illness. Key cost contributors were the treatment of moderate cases (31%), treatment of mild cases (29%) and initial testing (26%). Start-up and treatment of severe cases contributed 9%, and 5% to total costs respectively.

**Table 3:**
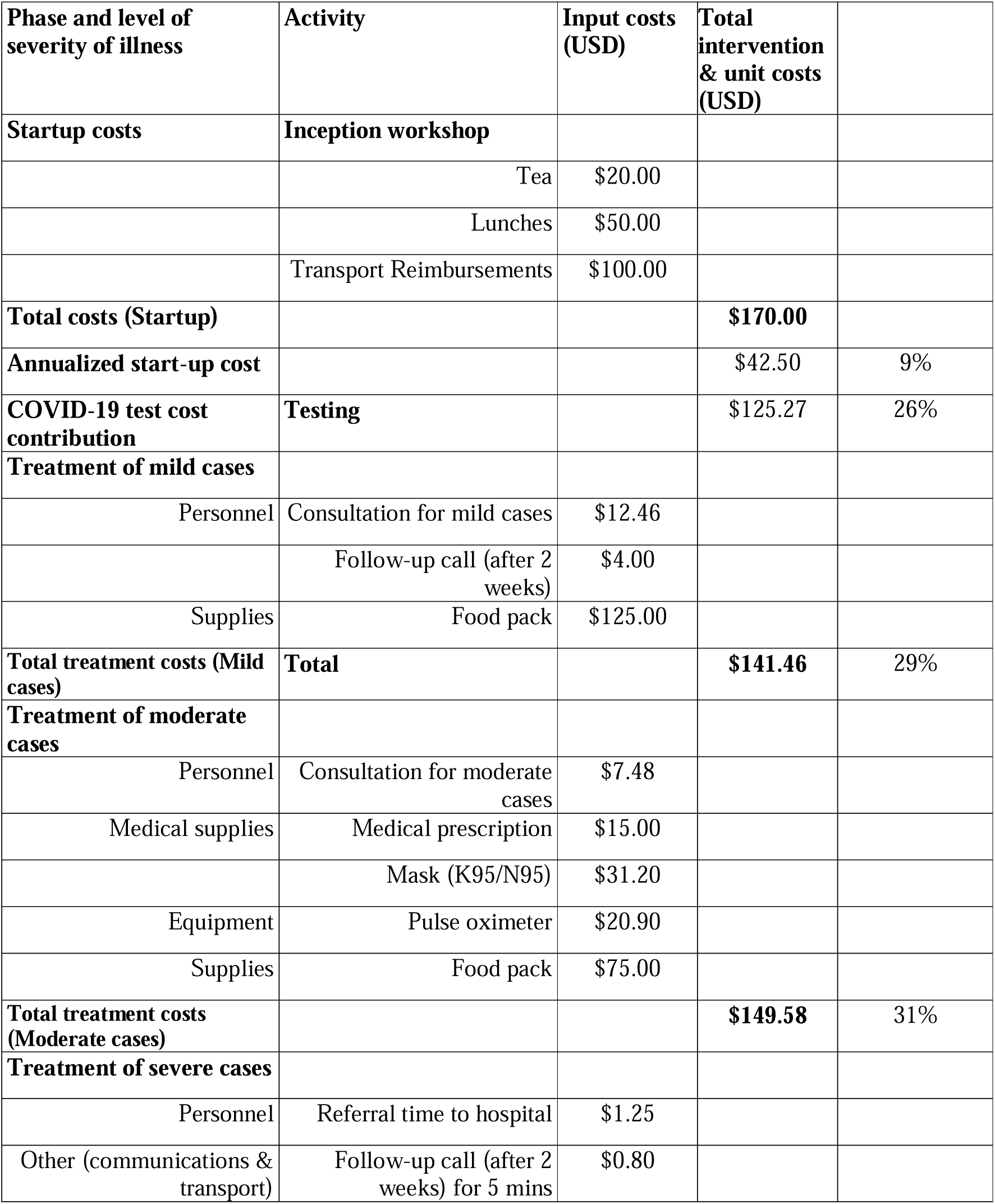
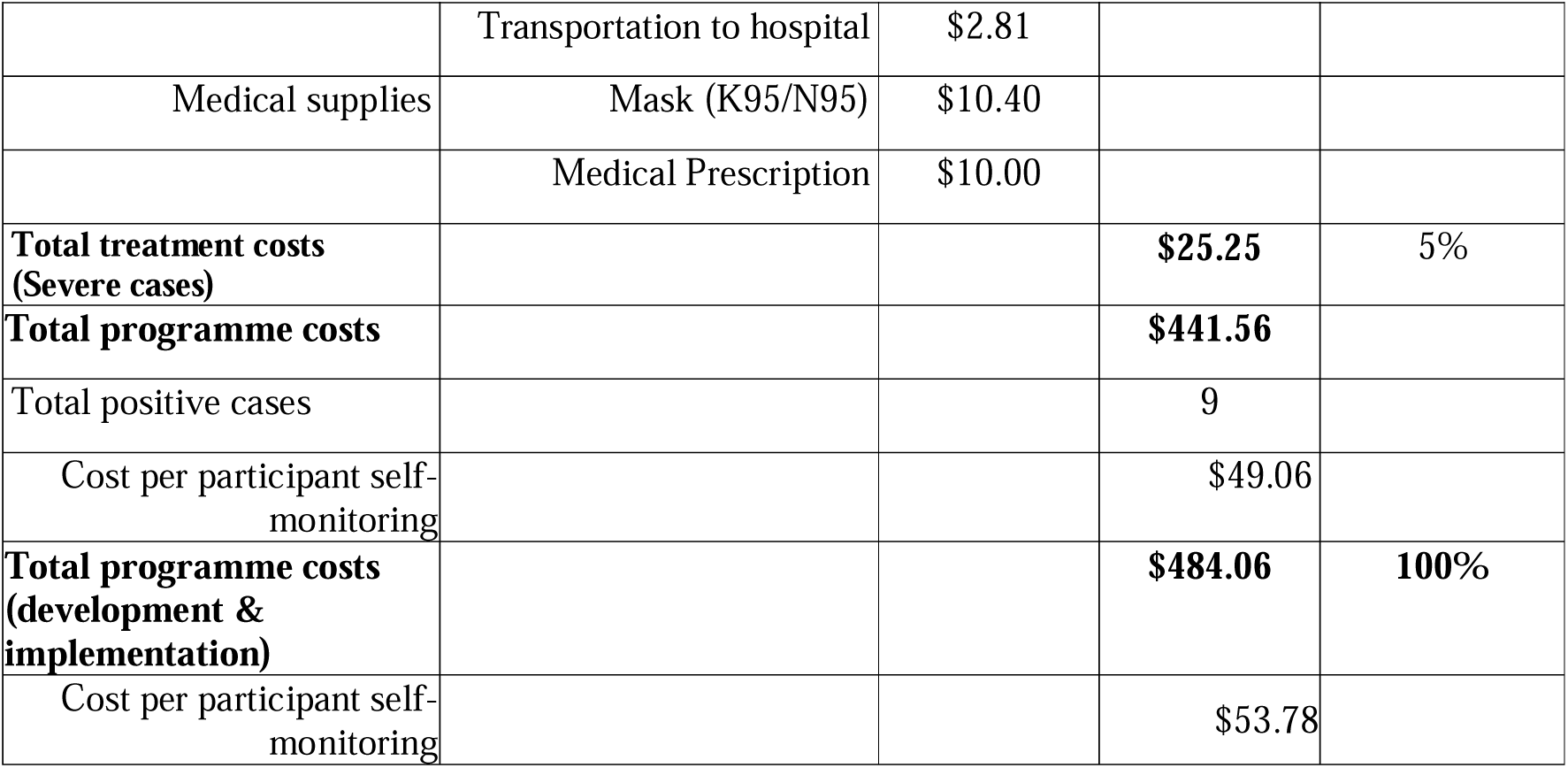
Programme and Unit Costs by Level of Severity of Illness.

| Phase and level of severity of illness | Activity | Input costs (USD) | Total intervention & unit costs (USD) |  |
| --- | --- | --- | --- | --- |
| **Startup costs** | **Inception workshop** |  |  |  |
| | Tea | \$20.00 | | |
| | Lunches | \$50.00 | | |
| | Transport Reimbursements | \$100.00 | | |
| **Total costs (Startup)** | | | **\$170.00** | |
| **Annualized start-up cost** | | | \$42.50 | 9% |
| **COVID-19 test cost contribution** | **Testing** | | \$125.27 | 26% |
| **Treatment of mild cases** |  |  |  |  |
| Personnel | Consultation for mild cases | \$12.46 | | |
| | Follow-up call (after 2 weeks) | \$4.00 | | |
| Supplies | Food pack | \$125.00 | | |
| **Total treatment costs (Mild cases)** | **Total** | | **\$141.46** | 29% |
| **Treatment of moderate cases** |  |  |  |  |
| Personnel | Consultation for moderate cases | \$7.48 | | |
| Medical supplies | Medical prescription | \$15.00 | | |
| | Mask (K95/N95) | \$31.20 | | |
| Equipment | Pulse oximeter | \$20.90 | | |
| Supplies | Food pack | \$75.00 | | |
| **Total treatment costs (Moderate cases)** | | | **\$149.58** | 31% |
| **Treatment of severe cases** |  |  |  |  |
| Personnel | Referral time to hospital | \$1.25 | | |
| Other (communications & transport) | Follow-up call (after 2 weeks) for 5 mins | \$0.80 | | |
| | Transportation to hospital | \$2.81 | | |
| Medical supplies | Mask (K95/N95) | \$10.40 | | |
| | Medical Prescription | \$10.00 | | |
| **Total treatment costs**<br>**(Severe cases)** | | | **\$25.25** | **5%** |
| **Total programme costs** | | | **\$441.56** | |
| Total positive cases |  |  | 9 |  |
| Cost per participant self-monitoring | | | \$49.06 | |
| **Total programme costs**<br>**(development & implementation)** | | | **\$484.06** | **100%** |
| Cost per participant self-monitoring | | | \$53.78 | |

## 4. Discussion

In this mixed-methods study conducted within the nationally scaled sex work program in Zimbabwe, we found that a co-developed intervention for supporting COVID-19 testing and community-based post-test self-monitoring with food support for sex workers was feasible and acceptable for sex workers and health workers. Sex workers were able to self-monitor their COVID-19 illness at home following instructions provided by health workers. Food support promoted adherence to self-isolation although vulnerability to other financial needs persisted. In depth interviews among health workers and sex workers revealed that stigma related to testing positive to COVID-19 prevented sex workers from seeking the health care they needed. The cost of intervention provision was $54 for each participant who had a positive diagnosis and conducted community-based self-monitoring.

The feasibility of community-based self-monitoring of COVID-19 illness by sex workers was facilitated by provision of understandable instructions, which are critical for ensuring that self-care is done correctly without harm to the patient/user [22]. Although instructions were largely understood, we found that there were instances of less-than-ideal comprehension which caused participants unnecessary anxiety. Testing/validation of the written instructions with intended users would have alleviated these challenges. Experiences from other self-care studies in Zimbabwe and Malawi highlight the importance of context-specific validation of instructions for use [23, 24].

Consistent with the literature [25], some sex workers failed to request the health care they needed because of fear of stigma related to testing positive for COVID-19. This adds to the stigma that sex workers already face related to their choice of profession and is a challenge that is encountered by sex workers in many other settings [26–28]. We add to the persistent calls to prevent stigma among sex workers so that there are no barriers to their uptake of services [28]. Specific recommendations for reducing COVID-19 stigma include avoidance of use of potentially stigmatizing language, use of media for disseminating anti-stigma messages, use of media to address COVID-19 myths and misinformation, ensuring public health messages empower individuals by suggesting safe ways to support those affected by COVID-19 and implementation of public health and structural interventions that target stigma [29–31]. These interventions should be complemented by program-level strategies aimed at enhancing individuals’ self-efficacy through education to assert right to health care despite reality of stigma [31].Programs can also borrow from successful stigma reduction activities in HIV, with appropriate tailoring for COVID-19. For example, in Zimbabwe, health education targeting HIV stigma reduction in schools was found effective in reducing stigma [32]. Weekly mass media radio sessions on HIV stigma and discussions centering on participants’ lived experiences were also found to reduce HIV stigma in Malawi [33].

Conditional incentives have been recognized in the literature as effective in promoting compliance with beneficial behaviours, particularly when linked to free access to basic health services [34]. In Tanzania, short-term food and cash assistance for food-insecure adults increased adherence to antiretroviral therapy (ART) by 85% in the cash group and 79% in the food group compared to standard care[35]. Similarly, food assistance recipients in Zambia demonstrated higher ART adherence than non-recipients[36]., Food packs reduced vulnerability to food insecurity and reportedly improved adherence to isolation by relieving financial pressure, which was also reported in other studies[7, 37, 38]. While food packs are a good solution, it is important to be aware of the limitations, such as residual vulnerabilities related to expenses such as housing, medical care, and education costs for children that are not covered by the food pack. Social protection programs were needed to address some of these vulnerabilities, and none were available for sex workers in Zimbabwe [39]. Many high-income countries were able to provide social protection through cash transfer, financial benefits and provision of emergency housing to sex workers [6, 8, 40]. It is crucial to include sex workers in social protection programs during pandemics. In countries like Nigeria and Uganda, where social protection programs existed but excluded sex workers, many continued sex work despite the risks of COVID-19 exposure [41–43]. This exclusion not only jeopardized their health but also highlighted the need for inclusive social protection that protect all vulnerable populations during pandemics.

Our cost analysis found that the cost per participant diagnosed with COVID-19 and self-monitoring at home was $54. This result compares favorably with findings of a modelling study that found very high costs of COVID-19 clinical management in low and middle income countries, with costs of clinical management of COVID-19 potentially as high as US$43.39 to US$75.57 per capita [44]. Of note, the same modelling study highlighted that scenarios with moderate or stringent physical distancing were associated with lower costs (US$1.10–US$1.32 per capita in the stringent physical distancing scenario). Our intervention promotes home-based monitoring and support for sex workers to adhere to isolation, hence we can expect wider cost savings from this intervention. Comparisons with specific programs in Africa also showed the intervention to have lower costs: daily per-patient costs for asymptomatic and mild-to-moderate COVID-19 disease patients under home-based care in Kenya (2020) were US$18.89 and US$18.99, respectively [45]. In Ghana the average cost of treating a COVID-19 patient from the health system perspective was US$282 for patients with mild/asymptomatic disease condition managed at home [46]. Implementation among larger numbers could bring the costs further down due to economies of scale.

The strengths of this study include using a mixed methods approach which allowed triangulation between the program data and the qualitative studies. We also include a cost analysis which can inform programs on development and implementation costs of this intervention. The intervention was co-developed with end-users and relevant stakeholders, which contributed to acceptability and feasibility. Limitations include the small number of sex workers who tested positive for COVID-19, although we were still able to explore in great depth how community-based self-monitoring was implemented. The small numbers of COVID-19 cases reflect the low disease burden that prevailed in the country during the study [47, 48]. From March 2022, the end of the fourth wave fueled by the Omicron variant, the country generally witnessed fewer positive cases, fewer hospitalizations and deaths [49]. The study positivity rate of 2.9% mirrors the low positivity rate in the general population.

## 5. Conclusion

A co-developed intervention for supporting adherence to COVID-19 isolation guidelines and community-based self-monitoring among sex workers in Zimbabwe was feasible and acceptable, with cost comparatively lower than other settings. In future, the intervention implementation can be optimized by reducing community stigma and further clarifying/simplifying instructions for self-monitoring.

## Data Availability

The data is available from authors upon reasonable request.

## Acknowledgements

We would also want to thank the reviewers who provided valuable insights to enhance the manuscript.

## AUTHORS CONTRIBUTION

**Conceptualisation**: Euphemia L Sibanda, Karin Hatzold, Elizabeth Corbett, Frances M Cowan

**Data curation**: Nancy Ruhode, Claudius Madanhire, Collin Mangenah, Itai Kabonga, Constance Watadzaushe

**Formal analysis**: Nancy Ruhode, Claudius Madanhire, Collin Mangenah, Itai Kabonga, Constance Watadzaushe

**Project administration**: Constance Watadzaushe, Itai Kabonga

**Supervision:** Euphemia L Sibanda, Karin Hatzold, Elizabeth Corbett, Yasmin Dunkley

**Writing of original drafts**: Itai Kabonga, Collin Mangenah

**Writing-reviewing and editing**: Itai Kabonga, Collin Mangenah, Constancia Watadzaushe, Claudius Madanhire, Nancy Ruhode, Yasmin Dunkley, Karin Hatzold, Elizabeth Corbett, Frances M Cowan, Euphemia L Sibanda

## SUPPORTING INFORMATION

S1: CeSHHAR COVID-19 screening form

S2: COVID-19 Symptom severity checklist

S3: Vulnerability assessment checklist

S4: 3ACP Codebook

S5: Oxygen saturation flyer

