## Supplementary material for "Feasibility, Acceptability, and Cost of Community-Based Self-monitoring among Sex Workers Testing Positive for COVID-19 in Zimbabwe: A Mixed-methods Study": S1

### CeSHHAR COVID-19 SCREENING FORM 3ACP

CeSHHAR COVID SCREENING FORM PLEASE REVIEW SOP BEFORE USING

*IF ONE IS HAVING DIFFICULTY BREATHING OR EXPERIENCING OTHER SEVERE SYMPTOMS, CALL 2019 TOLL FREE TO NOTIFY THE RAPID RESPONSE TEAM IMMEDIATELY.*

**Visit date**

2022-08-26

**Form completed by**

-

**Respondent is**

-

- ◯ Partner to SW
- ◯ New SW Client
- ◯ Old SW Client
- ◯ Visitor

**Please enter client's SISTERS number**

*X-X-000000*

#### Identifier information

**Please select name of site**

- ◯ BULAWAYO
- ◯ GWERU
- ◯ HARARE
- ◯ KAROI
- ◯ MASVINGO
- ◯ FORBES
- ◯ MUTARE
- ◯ BEITBRIDGE
- ◯ CHIRUNDU
- ◯ VICTORIA FALLS
- ◯ CHINHOYI
- ◯ KADOMA

**Please enter name of employee**

**Q1. Have you been vaccinated against COVID-19?**

- No
- Yes, one dose
- Yes, two doses
- Yes, two doses and booster

#### Onset of symptoms

**Q2. Do you have the following symptoms? (fever, new cough or difficulty breathing ) (or a combination of these symptoms)**

- Yes
- No

**Q3. What is the client's temperature reading?**

*If temperature reading is more than 37.5°C CALL 2019 TOLL FREE TO NOTIFY THE RAPID RESPONSE TEAM.*

**Q4. Is date of first symptom onset known?**

- Yes
- No

**Q5. Date of first symptom onset**

2022-08-26

#### Symptoms

**Q6. Do you have a fever?**

Yes No

**Q7. Do you have a new_cough?**

Yes No

**Q8. Are you having difficulty_breathing?**

Yes No

#### Symptoms

**Q9. Do you have muscle aches?**

Yes No

**Q10. Do you have a headache?**

- ◯ Yes ◯ No

**Q11. Do you have fatigue?**

- ◯ Yes ◯ No

**Q12. Do you have a sore throat?**

- ◯ Yes ◯ No

**Q13. Do you have runny nose?**

- ◯ Yes ◯ No

**Q14. Do you have diarrhoea?**

- ◯ Yes ◯ No

**Q15. Do you have loss of taste/smell?**

- ◯ Yes ◯ No

#### International human exposures in the days before symptom onset

**Q16. Have you or anyone you have been in contact with,travelled outside Zimbabwe in the last 14 days?**

- ◯ Yes ◯ No ◯ Don't know

**Q16a. Start of international travel.**

*Please write 01/01/2000 if cleint doesn't know the date*

yyyy-mm-dd

**Q16b. End of international travel.**

*Please write 01/01/2000 if cleint doesn't know the date*

yyyy-mm-dd

**Q17. Have you or anyone you have been in contact with been quarantined for COVID?**

- ◯ Yes ◯ No

**Q17a. Start of quarantine.**

*Please write 01/01/2000 if cleint doesn't know the date*

yyyy-mm-dd

**Q17b. End of quarantine.**

*Please write 01/01/2000 if cleint doesn't know the date*

yyyy-mm-dd

**Q18. Countries visited**

**Q19. Cities visited**

#### Case contact

**Q20. Does someone you are in close contact with have COVID-19 ( for example, someone in your household or workplace)**

◯ Yes ◯ No ◯ Don't know

**Q20a. Date of last contact.**

*Please write 01/01/2000 if cleint doesn't know the date*

2022-08-26

**Q21. Are you in close contact with a person who is sick with respiratory symptoms (for example fever, cough or difficulty breathing) who recently travelled outside Zimbabwe**

◯ Yes

◯ No

◯ Don't know

#### Offer COVID19 TEST

IF CLIENT ANSWERED YES TO ANY OF THESE QUESTIONS OFFER TESTING ACCORDING TO THE TESTING MODEL BEING IMPLEMENTED ON SITE

*IF CLIENT ANSWERED YES TO ANY OF THESE QUESTIONS OFFER TESTING ACCORDING TESTING MODEL BEING IMPLEMENTED ON SITE*

**Q22. Was client offered COVID testing?**

◯ Yes

◯ No

**Q22a. If no, why was testing not offered?**

◯ Trained staff who can do the test are not available

◯ Trained staff who can do the test are too busy

◯ Already tested elsewhere (State date dd/mmm/yyyy)

◯ Testing kits are not available in stock

◯ Other (specify)

**Q22a_Other. Specify other reason for not offering testing**

**Q23. Did client accept COVID testing?**

- ◯ Yes
- ◯ No

**Q23a. If no, reason for rejecting offer**

- ◯ No time
- ◯ Discomfort of sampling procedure
- ◯ Already tested elsewhere (State date dd/mmm/yyyy)
- ◯ Other (specify)

**Q23_Other. Specify other reason fo rejecting testing**

______________________________

**Q24. When was the covid test done?**

*Please write 01/01/2000 if cleint doesn't know the date*

2022-08-26

#### Form completion

**Q25. Form completed**

- ◯ Yes
- ◯ No

**Q26. Reason not completed**

- ◯ Missed
- ◯ Not attempted
- ◯ Not performed
- ◯ Refusal
- ◯ Other

**Q26a. Other reason not completed.**

______________________________
