## Supplementary material for "Feasibility, Acceptability, and Cost of Community-Based Self-monitoring among Sex Workers Testing Positive for COVID-19 in Zimbabwe: A Mixed-methods Study": S2

Clinicians will use this tool to assess and categorise patients who test positive for COVID-19. This assessment will determine next steps (e.g. referral to inpatient facility or recommendation to self-isolate or continue to self-isolate at home).

| **Illness category** | **Signs and symptoms** |
|---|---|
| **Mild** | No signs of respiratory distress |
| **Moderate** | Oxygen saturation is greater than 90% but less than or equal to 95%.<br>OR<br>**At least one of the following:** Fever >38 degrees Celsius, Respiratory rate 24 -30 and Pulse rate >120 and Systolic blood pressure <100<br>OR<br>Patient appears weak but can walk (based on clinical judgement) |
| **Severe** | Oxygen saturation is lower than 90%<br>OR<br>**All of the following:** Fever >38 degrees Celsius, Respiratory rate >30, Pulse rate >120 and Systolic blood pressure <100<br>OR<br>Patient is too weak to walk (based on clinical judgement) |
