## Supplementary material for "Feasibility, Acceptability, and Cost of Community-Based Self-monitoring among Sex Workers Testing Positive for COVID-19 in Zimbabwe: A Mixed-methods Study": S3

### VULNERABILITY ASSESSMENT CHECKLIST (To be used to determine clients that are vulnerable to severe disease and mortality due to COVID-19)

This is a starting version of the vulnerability checklist that needs to be revised in line with upcoming Ministry of Health guidelines

**Guidance for using this Vulnerability Checklist**

- This should be completed by Program Staff (Nurse or Outreach Worker)
- If a client has at least one of the conditions or factors listed in Appendix 1, treat them as vulnerable.

**Appendix 1: Conditions that make a client vulnerable**

1. Age more than 60 years (increasing with age).
2. Underlying noncommunicable diseases (NCDs):
    a. diabetes,
    b. hypertension,
    c. cardiac disease,
    d. chronic lung disease,
    e. cerebrovascular disease,
    f. dementia,
    g. mental disorders,
    h. chronic kidney disease,
    i. immunosuppression,
    j. HIV
    k. Obesity – score of 5 or higher for women and 6 or higher for men *(see Appendix 2 for determination)* and
    l. cancer
3. Other risk factors associated with higher risk include:
    a. smoking and
    b. In pregnancy,
        i. increasing maternal age,
        ii. high BMI, score of 5 or higher for women and 6 or higher for men *(see Appendix 2 for determination)*,
        iii. chronic conditions and
        iv. pregnancy specific conditions such as gestational diabetes and pre-eclampsia

**Classification into risk categories**

Low vulnerability – None of conditions that make a client vulnerable

High vulnerability – At least one of the conditions above in 1-3 above

#### 4. Acknowledgements:

This Risk Assessment has been adopted from the World health Organization Living Guidance for the Clinical management of COVID-19,version dated 23 November 2021.

#### Appendix 2: Self-determination of obesity

*Show the picture below to the participant, then ask this question*: The picture shows a scale of body sizes for men and women. Where would you place yourself on this scale?

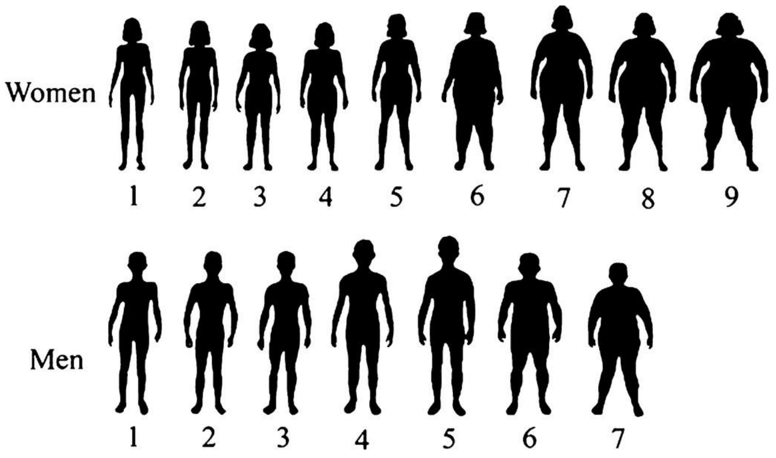
