## Supplementary material for "Feasibility, Acceptability, and Cost of Community-Based Self-monitoring among Sex Workers Testing Positive for COVID-19 in Zimbabwe: A Mixed-methods Study": S4

3ACP CODEBOOK

**Free nodes**

| **No.** | **Code** | **Description** |
| --- | --- | --- |
| 1. | Acceptance/adoption | This explores willingness of the individuals to do something e.g., to undergo testing for C19/or to deliver C19 testing and how contacts of the index patients reacted when approached with C19 self-test kits. How participants embrace new innovations. |
| 2. | Accessibility/convenience | Refer to the ease/difficult to access services/information this could include availability/reach/travel/affordability/location/time/overall service experience/ |
| 3. | Adequacy | Adequacy/Inadequacy of the various aspects responses and support systems e.g., facilities or patients lacking space/limited testing capacity/poor housing quality and inadequate conditions/understaffing issues. |
| 4. | Advice | Guidance and recommendations provided to individuals, and their sources/ to encourage the individual to act on something because of the perceived benefits. e.g., deciding to test for C19 because of advice from people e.g., doctor, friends, peers, or families. |
| 5. | Behaviours | This explores what the people do during a pandemic, when they hear about the pandemic, when they get symptoms or when exposed to someone with the C19, for example, what they do after testing and how the testing process/ result has modified their future behaviour. This also includes both positive and negative behaviours. |
| 6. | Beliefs | Perceptions that people hold regarding the pandemic: the etiology, transmission, prevention including testing influencing decisions to take actions during the pandemic. They can be positive or negative beliefs. |
| 7. | Benefits | Participant perceived/experienced positive outcomes/results from their engagement with health services. May include personal/public health benefits, long-term and short-term benefits. |
| 8. | Bereavement/death | Participants who lost friends and family during the pandemic. One of the impacts of C19, potential death or people dying. |
| 9. | Blame | Tendency of participants to assign fault or responsibility for something bad happening e.g., blaming those who test positive for C19 for the spread of the virus. |
| 10. | Burden | Perceived/experienced costs, inconvenience, negative impact associated with participants’ engagement with services during the pandemic. May include practical/emotional burdens, e.g., time and effort required to get tested/ combining usual role with the additional role in the intervention e.g., effects of combining the two roles (e.g., increasing/decreasing work burden), how it was like for them to manage the two tasks simultaneously, and how they managed the effects. |
| 11. | Capacity | Referring to the ability/inability of testing systems and infrastructure to meet the demands of the pandemic. Availability of testing supplies, number of trained providers to deliver the services, number of facilities available to the public. E.g., provider capacity to provide C19 testing services. |
| 12. | Children |  |
| 13. | Commitment | Willingness of individuals/government to support the engagement with services during the pandemic. E.g., political will, public support, following guidelines |
| 14. | Communication | Exchange of information and messaging related to the pandemic, may include the content of the messaging and ways in which it is delivered to different audiences. E.g., Learning about C19 through communication materials such as publicly displayed posters or images/social media |
| 15. | Comorbidities | Comorbidities are pre-existing medical conditions that can increase the severity or risk of complications from COVID-19. |
| 16. | Competence | Refers to the ability, knowledge, skills, and expertise required to perform a particular task or job effectively. E.g., How confident, or capable communities/providers feel doing what is required to meet objectives of this type of testing, how well they think they can meet the requirements of the intervention. How competent people felt about testing themselves for C19 to affect accurate and reliable results. |
| 17. | Compliance/adherence | The extent to which individuals are willing and able to participate in an intervention/a service. Willingness to follow SOPs/guidelines/prevention measures. Provider’s adherence to the C19 testing SOPs. And how consistently are participants able to follow these rule/guidelines/SOPs consistently. Participant’s view on future adoption and expansion of C19 testing. Participant’s compliance to C19 preventive measures. How this varied through the pandemic period, and the factors affecting the changes e.g., people being more adherent in the early stages when there more cases and deaths and they felt at risk, or because the enforcement was stricter. |
| 18. | Confidentiality | Need to protect the privacy of the individuals participating in an intervention or engaging with a service. Protecting individuals’ personal information and other identifiers. Safeguarding of the test results. Including standard that are put in place to ensure that privacy and confidentiality of participants’ information is protected. Saying C19 self-testing was better than provider-delivered testing because was done in private and was therefore less stigmatizing. |
| 19. | Contact | Participants who tested because they had exposure to positive cases. Who their contacts were, the method they used to approach them with C19 self-test kits, and where they met them. |
| 20. | Cope | Ability to manage emotional/psychological/practical challenges associated with the pandemic and the strategies that were enforced |
| 21. | Counselling | Psychological support and guidance to individuals who are considering, for example, testing. It involves explaining the testing process, expected outcome and post-test action to participants by provider prior to testing |
| 22. | Decision | Process of making a choice or taking action. Decisions on testing/adherence/adoption, the things that influence them to decide to test. … it can be suspected C19 related symptoms, desire to access treatment. Failure to test for C19 because they already tested previously. Saying they decided or accepted to test for C19 for no particular reason, or because they just felt good about doing it, or they just felt an ardent desire to do it. |
| 23. | Delivery | Providing services to participants in a manner that is accessible, convenient and efficient and the strategies being used. May also include appointment scheduling, wait times, speed of the delivering the service. |
| 24. | Denial/othering | State of being resistant to do/feel something E.g., refusing to test for C19 when an individual is experiencing symptoms. Refusing to take a C19 test because they don’t believe they would be infected with the virus, thinking only certain people might have the virus |
| 25. | Disclosure | Process of informing others about their health status. When, to whom and how it happens and why. |
| 26. | Discrimination/stigma | Unfair treatment/stigmatization of individuals based on their health status, occupation. Discrimination associated with disclosure, especially if positive C19 status was made known to people that were non-relations or not close friends. May be self/social, perceived/experienced. |
| 27. | Employment |  |
| 28. | Encouragement | Promoting and supporting others, providing information about the benefits of testing, addressing concerns and fears that people may have, includes sources. Issues that encourage/discourage participants to test/deliver testing |
| 29. | Enrolment/screening | Descriptions of how the research team approached and invited them to participate or test. This describes how they were approached and offered C19 testing services by the study, and how they felt about the approach. |
| 30. | Experience | Provider work experience associated with current role in C19 testing services. Participant’s positive/negative previous testing experience leading to testing refusal. What involvement they have had in C19 service provision prior to this current testing service e.g., diagnostics, treatment, prevention, etc. |
| 31. | Face mask | Participants’s knowledge, experience with using face mask as a C19 preventive measure. One of the C19 preventive measures, including explanations of using the mask correctly. |
| 32. | Facility | Physical locations where services are being delivered, description of the structures. |
| 33. | Falsification | Perceived drawback of C19 self-testing, possibility of people lying about using the kit when they did not, or telling a false test result |
| 34. | Familiarity | Acquaintance with or having prior knowledge/experience with something, e.g., self-testing, signs and symptoms. E.g., Inability to tell whether the symptoms they are having are C19 due to similarities between C19 symptoms and those of other conditions, which meant people were unsure whether or not they needed a C19 test. |
| 35. | Fate/fatalism | The belief that something is predetermined and beyond the control of individuals/healthcare providers. Feeling that taking a C19 test was only a matter of time as this was always going to happen, indicating a sense that predetermined natural events dictated their choice to test. |
| 36. | Fatigue | Feeling tired/exhausted from the ongoing efforts of complying to protocols. Participants refusing to continue serial testing for 12 weeks during workplace screening. One of the C19 symptoms. |
| 37. | Fear | Apprehensions that people may experience about the pandemic. Participant’s fear of being positive leading to testing refusal. Fear impacting how people respond to different aspects of the pandemic. E.g. fearing death caused health-promoting behaviours e.g. people were more willing to vaccinate against C19 when cases and deaths were more prevalent. Fear of complications also dissuaded people from vaccinating. Having fears about getting infected (especially in the early days of the pandemic) and this aiding compliance as people were more willing to use the preventive measures |
| 38. | Finance | Anything to do with money, availability or lack of it. Includes poverty, fees etc. people losing income from work/business. |
| 39. | Frequency | How often testing is required/recommended/availability of a service. How often people should test for C19. How many times they tested, either through the provider-delivered method or the self-test approach. Includes participants’ views on frequency. |
| 40. | Hand washing | One of the methods of preventing C19, including descriptions of handwashing is supposed to be done (how and how often). Participants experience with hand washing as a C19 preventive measure. |
| 41. | Hardship | Economic, physical and psychological impact of C19 experienced by participants during the pandemic |
| 42. | Health talk | Any communication related to health and healthcare to increase knowledge and awareness of health issues, promote health behaviours and habits and improve overall health outcomes. |
| 43. | Helplessness | Feeling powerless or lack of control, deciding or accepting to take a C19 test (primarily) because they were sick and looking for treatment, indicating a sense of powerlessness in which accepting to test is seen as a gateway to treatment. |
| 44. | Illness | Poor health/disease. Whether or not they have had C19 previously |
| 45. | Information | Any facts/or not conveyed from one person to another, it can be accurate/inaccurate, complete or incomplete. Also includes sources of information on different aspects of the C19 pandemic e.g. where they learnt about vaccine, testing, etc |
| 46. | Integration | How C19 testing service providers integrate C19 testing into their daily health care service provision routines |
| 47. | Interpretation | Understanding the meaning of the test results, reliability and accuracy of the testing method. |
| 48. | Isolation | One of the steps they followed in order to avoid transmitting the virus after testing positive. How they/others felt about isolation/and the experience of it. |
| 49. | Knowledge | Understanding of or insights that people have about the pandemic. Participant knowledge about C19, transmission, risk factors, symptoms, treatment, and prevention. Access to C19 services. Participant literacy level and ability to use the self-testing kit unassisted. Participants’ understanding of the purpose of the 3ACP intervention. |
| 50. | Lockdown | Closing no-essential businesses, restricting gatherings, requiring people to stay at home except for essential activities, all imposed by governments or public health authorities to limit the spread of the virus. Participants shares their experience with the prevention of movement during the C19 pandemic and the impact in their lives |
| 51. | Mandates | Government requirement/instruction to help control the spread of the virus. E.g., testing/vaccination as a condition for international travel. |
| 52. | Motivation | What encouraged/discouraged the subjects to take the test. Both from internal/external sources. |
| 53. | Novelty | Newness of the testing procedure |
| 54. | Peer |  |
| 55. | Preference/choice | Participant choice of either self-test or professional use when offered Ag RDT test kits. Reasons for testing preference |
| 56. | Procedure | Description how test was done, how participants were approached, preparing clients for testing, how the sample was collected, and test result presented. How they felt about it. |
| 57. | Provider |  |
| 58. | Referral | Directing/recommending a person to seek further medical care, treatment or evaluation by another healthcare provider. Patients that tested positive for C19 saying they were told to go and buy drugs from a pharmacy/drugstore. |
| 59. | Relevance | How important they feel the testing intervention is to their target population, general population, or the country, health system. What the needs of the target population are, how these needs are met, and whether any needs are not being met, why they are not met, what can be done to ensure they are met. |
| 60. | Result/outcome | The test results of participants who did C19 test. How they knew that they had C19 e.g., how were they told… and what pushed the decision to test, symptoms they had, etc. How did they react to being diagnosed with C19. Test results of the contacts when they self-tested.This should also capture the reaction of those whom they told about their positive/negative status. |
| 61. | Risk | Risk of contracting C19, risk of false negatives/positives or risks associated with testing procedure (discomfort/pain). Risk for self or wider community/country. Increasing/decreasing risk |
| 62. | Role model | Volunteering to take a C19 test out of free will so as to inspire others to imitate their behaviour and promote community uptake of C19 testing. |
| 63. | Rumours | False/unverified information about C19 that is being spread through social networks or other forms of communication. Including sources. |
| 64. | Safety | Perceptions/experience that the intervention/services are free from harm, danger, or risk. Includes appropriateness, availability of protective equipment, competencies, and adherence. |
| 65. | Self | An individual’s own being, identify. Being concerned about their own health as a reason for accepting to test or self-test for C19 |
| 66. | Sensitisation | Informing individuals about the importance and benefits of service/intervention testing, raising awareness and knowledge on C19, testing methods available, importance of testing, and how to get tested. |
| 67. | Severity | Perceptions of how serious C19 is or can be (if people don’t take precautions). Participant’s views about the severity of C19 in their area/country during, previously and currently. |
| 68. | Side effects | Adverse/unwanted symptoms or reactions that may occur as a result treatment/vaccines |
| 69. | Social/physical distancing | Practice of maintaining a safe distance from others to minimise the spread of C19. Time spent near others and avoiding physical contact. Participants experience with social or physical distancing as a C19 preventive measure. |
| 70. | Solidarity | Families or neighbours coming and working together to deal with the effects of the pandemic. |
| 71. | Stress | Emotional strain that individuals may increase related to C19. Stress from getting a positive result, impact of C19 on their health or the health of others, anxiety due to an increased economic burden (shortage of money for food, money for school fees, etc), caused by sickness with C19 and the need to quarantine, or national stay-at-home orders, which meant they were unable to work and provide for themselves and their families. |
| 72. | Support | Types of emotional, social, economic and clinical support; provided to clients by providers pre and post testing, words of encouragement from friends (if they disclose). This is about if they were supported to approach and deliver self-test kits to their contacts. How they felt about providers asking them to test for C19 e.g., seeing the doctor’s recommendation as an expression of kindness and wishing them well. Showing sympathy and concern e.g., spouse wanting to stay close sharing the same isolation room with them despite the risk or praying for their recovery. |
| 73. | Symptoms | Participant’s knowledge of the symptoms of C19. Symptoms they experienced when they had C19 e.g., were they sick? Having symptoms prompting them to come to the clinic for examination. |
| 74. | Test | All information related to testing for C19. Testing procedure, participants showing confidence in the COVID-19 testing process as a result of having been previously tested. Participants completing 12 weeks C19 screening testing at the workplace. Collecting self-test kits for contacts and for themselves. Index patients using some of the self-test kits collected for their contacts to self-test themselves to see if they still have the virus. Retesting, self-testing, provider-delivered testing |
| 75. | Training | Trainings attended by C19 testing service providers to enhance C19 testing service provision. |
| 76. | Transmission | Participant knowledge of the mode of transmission of C19. Various ways through which participants feel C19 can be transmitted. |
| 77. | Treatment | Availability and effectiveness of treatment. Participant who thinks seeking treatment is one of the immediate action to take when outcome of a COVID-19 test is positive (behaviour) |
| 78. | Trust | Extent to which individuals believe/not that there is C19, reliability/accuracy/safety of the testing/vaccine process. Trust in the information on C19, trust in others/systems. Lack of trust in the government and Health care workers in COVID-19 and the COVID-19 vaccine. E.g., refusing to test for C19 because of disbelief in healthcare workers, suspecting that providers were being paid and used to get people tested |
| 79. | Vaccination | Participants’ vaccination status including how many times they vaccinated, which vaccine and their views about vaccines. |
| 80. | Workplace | An organized setting either government or private with employees. Where they work or do their business |
| 81. | Worry | Concern/unease about a situation/potential problem/others. Worrying that they might infect others (family members, friends) and taking a test to know status and be better informed about what to do to ensure safety of others. |

**Attributes**

- Age
- Highest level of education
- Occupation
- Gender
- Marital status
- Years in a role
- Years in an organization

**Sets/folders**

- Use cases

**Categories**

1. **Access and availability:**
   a. Accessibility/convenience
   b. Adequacy
   c. Capacity
   d. Delivery
   e. Enrolment/screening
   f. Referral
   g. Relevance
2. **Attitudes and perceptions**
   a. Beliefs
   b. Benefits
   c. Blame
   d. Commitment
   e. Denial/othering
   f. Discrimination/stigma
   g. Fate/fatalism
   h. Novelty
   i. Preference/choice
   j. Risk
   k. Trust
3. **Behavioural response:**
   a. Acceptance/adoption
   b. Behaviours
   c. Competence
   d. Compliance/adherence
   e. Cope
   f. Decision
   g. Face mask
   h. Frequency
   i. Hand washing
   j. Integration
   k. Isolation
   l. Knowledge
   m. Lockdown
   n. Mandates
   o. Social/physical distancing
   p. Test
   q. Transmission
   r. Vaccination
4. **Communication and knowledge/information:**
   a. Advice
   b. Communication
   c. Disclosure
   d. Experience
   e. Falsification
   f. Familiarity
   g. Health talk
   h. Information
   i. Interpretation
   j. Rumours
   k. Sensitisation
5. **Emotional/mental well-being**
   a. Burden
   b. Denial/othering
   c. Encouragement
   d. Fear
   e. Hardship
   f. Helplessness
   g. Stress
   h. Worry
6. **Finance**
7. **Events**
   a. Illness
   b. Bereavement/death
8. **Treatment and care:**
   a. Health condition/symptoms
      i. Comorbidities
      ii. Fatigue
      iii. Symptoms
      iv. Severity
      v. Side effects
   b. Confidentiality
   c. Healthcare services
      i. Counselling
      ii. Procedure
      iii. Result/outcome
      iv. Treatment
      v. Safety
   d. Patient support
      i. Solidarity
      ii. Support
9. **People**
   a. Peer
   b. Provider
   c. Self
   d. Children
   e. Contact
10. **Places**
    a. Facility
    b. Workplace
