## Supplementary material for "Feasibility, Acceptability, and Cost of Community-Based Self-monitoring among Sex Workers Testing Positive for COVID-19 in Zimbabwe: A Mixed-methods Study": S5

**A programme for providing COVID 19 testing services at CeSHHAR clinics**

**Oxygen Saturation flyer for sex workers**

The pulse Oximeter is an instrument/ device that is used to measure the amount of oxygen in your blood. It can be used to determine whether your COVID-19 illness has worsened to a level that requires you to go to hospital.

- Please note that the pulse oximeter is only one way of determining progression of COVID-19 illness. Please contact the peer educator or outreach worker if you have any of the following **"danger signs"** regardless of pulse oximeter readings:
    - muddled or confused thinking
    - difficulty breathing
    - fast breathing
    - chest pain,
    - difficulty walking

**Below, please find instructions on how to use the pulse oximeter:**

- We recommend that you use the pulse oximeter twice a day during the period when you are feeling ill

**How to use the pulse oximeter**

Before using the Oximeter, clean the rubber hole with alcohol or sanitiser. The rubber hole is found below the blue surface. Clean your finger before and after the test.

- Place your finger into the rubber hole of the Oximeter, with your nail surface upward.
- Switch on the Oximeter by pressing the white button on top of the Oximeter for 3 seconds
- The Oximeter will start recording.
- **DO NOT** shake your finger or your body when the Oximeter is working
- Read the number showing in yellow on the screen and write it down. This is your oximeter reading
- When you remove your finger, the device will automatically turn off

**This is how you should interpret the pulse oximeter readings:**

**Oximeter reading of 95% and over**

- This is a normal level of oxygen in your blood and doesn't need to be repeated.
    - *But please still report promptly any "danger signs" listed above*

**Oximeter reading of under 95%**

- This is a low reading – repeat the test to confirm the result and report it straight away
    - Confirm - sit down comfortably and repeat the reading with your hand still.

        - You don’t need to take any action if the second reading is normal (95% or higher)
    - A second reading below 95% needs to be reported to a Microplanner or Outreach worker using the phone number you have. The Microplanner or outreach worker will in turn alert the program nurse.
- A program nurse will telephone you to discuss your symptoms with you and determine whether you should be referred to the hospital or not.
- If you should go to the hospital, the nurse will order an ambulance to transport you to the hospital or ensure that you are transported to the hospital according to mechanisms employed by the local COVID-19 rapid response task force team.
- Treatment of COVID-19 patients is provided for free at public hospitals.

**Oximeter reading of 90 % - confirmed on 2 readings**

- If your first and second readings confirm oxygen levels of 90% or below, you need to go into hospital
- You must immediately report by phone to your assigned Microplanner or Outreach worker who will in turn alert program staff. Microplanners and Outreach workers have been trained to treat readings of 90% and below with urgency, so be ready to go to hospital as soon as you have placed the report.
- Program staff will order an ambulance to transport you to the hospital or ensure that you are transported to the hospital according to mechanisms employed by Ministry of Health.
